# MultiSCRIPT-Cycle 1- A Pragmatic trial embedded within the Swiss Multiple Sclerosis Cohort (SMSC) on neurofilament light chain monitoring to inform personalized treatment decisions in Multiple Sclerosis: a study protocol for a randomized clinical trial

**DOI:** 10.1101/2024.03.22.24304720

**Authors:** Perrine Janiaud, Chiara Zecca, Anke Salmen, Pascal Benkert, Sabine Schädelin, Annette Orleth, Lilian Demuth, Aleksandra Maleska Maceski, Cristina Granziera, Johanna Oechtering, David Leppert, Tobias Derfuss, Lutz Achtnichts, Oliver Findling, Patrick Roth, Patrice Lalive, Marjolaine Uginet, Stefanie Müller, Caroline Pot, Robert Hoepner, Giulio Disanto, Claudio Gobbi, Leila Rooshenas, Matthias Schwenkglenks, Mark J Lambiris, Ludwig Kappos, Jens Kuhle, Özgür Yaldizli, Lars G Hemkens

**Affiliations:** Research Center for Clinical Neuroimmunology and Neuroscience Basel (RC2NB), University Hospital Basel, University of Basel, Basel, Switzerland; Neurology clinic Lugano, MS center, Neurocenter of Southern Switzerland, Lugano, Switzerland; Faculty of Biomedical Sciences, Università della Svizzera Italiana (USI), Lugano, Switzerland; Department of Neurology, Ruhr-University Bochum, St. Josef-Hospital, Bochum, Germany; Department of Clinical Research, University Hospital Basel, University of Basel, Basel, Switzerland; Neurologic Clinic and Policlinic, MS Centre, University Hospital Basel, Basel, Switzerland; Department of Neurology, University Hospital Basel, University of Basel, Basel, Switzerland; Translational Imaging in Neurology Basel, Department of Medicine and Biomedical Engineering, University Hospital Basel, University of Basel, Basel, Switzerland; Department of Neurology, Cantonal Hospital Aarau, Aarau, Switzerland; Department of Neurology, University Hospital Zurich and University of Zurich, Zurich, Switzerland; Division of Neurology, Department of Clinical Neurosciences, Faculty of Medicine, Geneva University Hospitals, Geneva, Switzerland; Department of Neurology, Cantonal Hospital St. Gallen, St. Gallen, Switzerland; Service of Neurology, Department of Clinical Neurosciences, Lausanne University Hospital (CHUV) and University of Lausanne, Lausanne, Switzerland; Department of Neurology, Bern University Hospital and University of Bern, Bern, Switzerland; Bristol Population Health Science Institute, University of Bristol, Bristol, UK; Health Economics Facility, Department of Public Health, University of Basel, Basel, Switzerland; Institute of Pharmaceutical Medicine (ECPM), University of Basel, Basel, Switzerland

## Abstract

**Background:** Treatment decisions for persons with relapsing-remitting multiple sclerosis (RRMS) rely on clinical and radiological disease activity, the benefit-harm profile of drug therapy, and preferences of patients and physicians. However, there is limited evidence to support evidence-based personalized decision-making on how to adapt disease modifying therapies treatments targeting no evidence of disease activity, while achieving better patient-relevant outcomes, fewer adverse events and improved care. Serum neurofilament light chain (sNfL) is a sensitive measure of disease activity that captures and prognosticates disease worsening in RRMS. sNfL might therefore be instrumental for a patient-tailored treatment adaptation. We aim to assess whether 6-monthly sNfL monitoring in addition to usual care improves patient-relevant outcomes compared to usual care alone.

**Methods:** Pragmatic multicenter, 1:1 randomized, platform trial embedded in the Swiss MS Cohort (SMSC). All patients with RRMS in the SMSC for ≥1 year are eligible. We plan to include 915 patients with RRMS, randomly allocated to two groups with different care strategies, one of them new (group A), one of them usual care (group B). In group A, 6-monthly monitoring of sNfL will together with information on relapses, disability and magnetic resonance imaging (MRI) inform personalized treatment decisions (e.g., escalation or de-escalation) supported by pre-specified algorithms. In group B, patients will receive usual care with their usual 6- or 12- monthly visits. Two primary outcomes will be used: 1) evidence of disease activity (EDA3: occurrence of relapses, disability worsening, or MRI activity) and 2) quality of life (MQoL-54) using 24-month follow-up. The new treatment strategy with sNfL will be considered superior to usual care if either more patients have no EDA3, or their health-related quality of life increases. Data collection will be embedded within the SMSC using established trial-level quality procedures.

**Discussion:** MultiSCRIPT aims to be a platform where research and care are optimally combined to generate evidence to inform personalized decision-making in usual care. This approach aims to foster better personalized treatment and care strategies, at low cost and with rapid translation to clinical practice.

**Trial registration:** NCT06095271

## Introduction

Multiple sclerosis (MS) is an inflammatory, demyelinating and neurodegenerative disease of the central nervous system, typically affecting persons in early adulthood, and is a leading cause of non-traumatic disability in young adults (1). MS presents heterogeneous courses of the disease, the most common form being relapsing-remitting MS (RRMS), with high variability in symptoms and treatment responses. There are currently over 20 disease modifying therapies (DMTs) that have been approved but all have diverse benefits, harms and burdens (2). High efficacy DMTs such as for example natalizumab (3), alemtuzumab (4) and ocrelizumab (5) which are given as infusions, ranging from every 4 weeks to yearly, lead to an almost complete suppression of acute disease activity. However, such DMTs inevitably inhibit the natural immune response putting patients at risk for harm due to, for example viral or bacterial infections (6).

As more potent DMTs are being developed (7) one should also explore the optimization of currently available DMTs (8). Personalized treatment strategies for persons with MS are urgently needed (8,9) to treat patients as little as possible but as much as necessary and at the right time (10). Practically, this means ensuring no evidence of disease activity, while achieving better patient-relevant outcomes such as, for example, improved quality of life, fewer adverse events, and improved care. Such an approach requires detailed information on disease activity and treatment response. Currently, in usual care this includes information on relapses, new/enlarging T2 weighted (w) MRI lesions or T1w contrast enhancing lesions, and confirmed disability worsening. MRIs are time and resources consuming, and occurrence of disease worsening despite stable standard MRI features is well documented (11). There is an urgent need for a body fluid biomarker (9) allowing for reliable and rapid detection of disease activity leading to prompt treatment escalation or as important additional surveillance to ensure disease stability during treatment de-escalation.

Serum neurofilament light chain (sNfL) has emerged as a promising fluid biomarker reflecting neuro-axonal damage in MS and correlating with disease activity and severity (12–15). Recent advancements in immunoassay technology allow for sensitive detection of subtle sNfL level increases in serum samples (16). sNfL levels are associated with future MS disease activity, disability worsening, MRI activity, and treatment response (17–22). When added to clinical assessments (i.e., relapses and disability worsening assessment) and conventional MRI, sNfL increases sensitivity in detecting disease activity and worsening in disability score (14). However, while sNfL shows potential for personalized treatment decisions, its routine use in clinical care is not widespread or recommended in major clinical guidelines. To best of our knowledge, there are currently no planned or ongoing randomized controlled trials assessing the clinical usefulness of sNfL in MS therapy monitoring in clinical practice.

### Objectives

The primary objective of this trial is to assess whether a treatment strategy including sNfL monitoring improves patient-relevant outcomes and care of patients with RRMS by either increasing the proportion of patients with no evidence of disease activity or by improving patients’ health-related quality of life. We assume that introducing a 6-monthly monitoring of sNfL within SMSC usual care will inform more personalized treatment decisions and result in either:

1. Better quality of life for patients with MS through biomarker (i.e., sNfL) guided de-escalation by reducing treatment burden and risk for side effects associated with highly effective DMT, or
2. Lower disease activity by early and/or more sensitive biomarker-guided escalation of DMT.

Secondary objectives include assessing if a treatment strategy including 6-monthly monitoring is associated with a decrease in the proportion of patients with relapses, disability worsening (assessed using Expanded Disability Status Scale (EDSS) scores) and/or MRI activity (i.e., new/enlarging T2 weighted (w) lesions or T1w contrast enhancing lesions). We also aim to assess if specific patient subgroups benefit from 6-monthly sNfL monitoring, i.e., patients at higher risk in comparison to patients at lower risk for future disease activity. We further aim to evaluate the economic implications of monitoring sNfL in terms of direct and indirect RRMS-related costs, quality-adjusted life-time, and incremental cost-effectiveness.

## Methods

MultiSCRIPT-Cycle 1 is a pragmatic multicenter, 1:1 randomized, trial embedded in the SMSC to compare a new treatment strategy including sNfL monitoring in addition to SMSC usual care compared with SMSC usual care alone. Patients are randomized in a 1:1 ratio to two groups with these different treatment strategies, one of them new (group A) and one of them usual care (group B).

MultiSCRIPT-Cycle 1 is a pragmatic trial that aims to provide evidence closely reflecting what happens in routine care (PRagmatic Explanatory Continuum Indicator Summary (PRECIS-2) (23) available in Appendix 1). It is patient-centered, and clearly focused on real-world decision making. It is embedded in the existing SMSC data structure and is based on routinely collected data from well-established and field-tested processes with central quality controls.

### Trial Setting

MultiSCRIPT-Cycle 1 is fully embedded in the Swiss MS Cohort (SMSC), leveraging the existing research infrastructure and processes. The SMSC (NCT02433028 – BASEC ID: PB_2016-01171) is a prospective multicenter cohort study performed across eight Swiss academic medical centers (the University Hospitals of Basel, Berne, Geneva, Lausanne and Zurich, and the Cantonal Hospitals of Aarau, Lugano and St. Gallen).

Usual care within the SMSC consists of 6 or 12-monthly clinical visits, including routine assessment of relapses and disability status (measured using the Expanded Disability Status Scale; EDSS), a blood draw (mandatory within SMSC usual care) and may include MRI at the discretion of the treating physician and patient’s preferences (facultative but it is routine for persons with MS to get yearly MRI to assess new/enlarging T2w lesions and/or T1w contrast enhancing lesions on cranial and/or spinal MRIs) (24). MRI protocols are standardized and aligned across centers. The centers scan the patients by default always at the same scanner with the same scanning parameters (e.g. head position in the scanner) to ensure maximum of comparability between the scans.

The SMSC 6 or 12-monthly schedule is at the discretion of the physician and patient’s preferences, and may vary over time. The SMSC collates routinely collected data (i.e., data not collected for the purpose of research) into a standardized and unique database and all blood samples collected as part of the SMSC usual care are biobanked.

### Eligibility criteria

We include all patients who have been diagnosed with RRMS according to the most recent McDonald criteria (2017) (25) for at least a year and have already consented to take part in the SMSC. Patients who are included (or planned to be included) in another DMT trial are excluded as they are (or will) most likely not follow the SMSC usual care.

The eligibility criterion that participants must have been diagnosed with RRMS for at least a year is used because it frequently takes up to a minimum of a year of treatment before a DMT adaptation may be considered. We are explicitly not excluding pregnant women from MultiSCRIPT-Cycle 1, or patients with specific conditions or concomitant diseases. We are aiming to generate evidence for all patients, including vulnerable populations, for which the tested intervention may be used in real-world care settings.

Patients who are not participating in MultiSCRIPT Cycle 1 but are in the SMSC will serve as external control subjects.

### Primary Outcomes

The two independent primary outcomes are (1) EDA3 (evidence of disease activity) and (2) quality of life using the Multiple Sclerosis Quality of Life (MSQoL)-54 instrument. Both primary outcomes will be assessed using 24 months follow-up data (i.e., 24 months since randomization).

EDA3 is defined as the occurrence of a relapse as defined in the McDonald criteria (25), confirmed disability worsening defined as an EDSS increase of ≥1.5 steps if baseline EDSS was 0, ≥1.0 step if baseline EDSS 1.0 to 5.5 and 0.5 steps if baseline EDSS >5.5 (11), or new/enlarging T2w lesions compared to the last MRI or T1w contrast enhancing lesions based on local MRI readings. EDA3 has better predictive value of disease worsening compared to taking its components individually (26) and NEDA3 is regarded as the most adequate indicator of treatment response (27).

The MSQoL-54 Instrument is an extension of the well-established Short Form-36 (SF-36) specifically for MS patients. It is a validated instrument with an adequate test-retest reliability, construct validity and internal consistency (28,29). MSQoL-54 is a structured self-reported questionnaire including 54 items generating 12 subscales with two summary scores, the physical health composite summary and the mental health composite summary (28). For the primary outcome, we will use the sum of both composite summaries as a total score (30).

### Secondary outcomes

The secondary outcomes include assessments of EDA3 and MSQoL-54 at 12 months. The individual components of EDA3 will also be assessed separately including relapses, disability worsening measured by EDSS, and new/enlarging T2w lesions and T1w contrast enhancing lesions based on local MRI reading and on centralized MRI readings. Similarly, the individual summary scores of MSQoL-54 will also be assessed separately. The amount of immunosuppressive/ immunomodulatory drug treatment (or DMTs) will be monitored. Quality of life will be further assessed using EQ-5D-5 and SF-36. All secondary outcomes will be assessed using 12- and 24-month follow-up data (i.e., 12 and 24 months since randomization).

For the health economical evaluation, we will assess health-related quality of life measured with the EQ-5D-5L, quality-adjusted life years, professional activity status and change, indirect costs, and direct medical costs based on healthcare utilization (e.g., hospitalizations).

To better understand treatment pathways and clinical decision making, we will collect information on treatment changes (e.g., how many patients were escalated or de-escalated) and the reason for treatment change.

### Harm outcomes

Any serious adverse events related to the key intervention in the new strategy (i.e., blood draw for sNfL measurement) will be monitored and collected until the end of the study conduct at 42 months of follow-up.

In addition, the following harm indicators will be assessed during the safety interim analysis: mortality, harms related to immunosuppression (e.g., relevant infections), occurrence of relapses and/or disability worsening in patients previously stable.

All follow-up data on SAEs and indicators of harms available at the database closure will be used for the safety analysis.

### New care strategy (group A)

At randomization, patients are allocated to a new treatment strategy (group A) with 6-monthly sNfL monitoring in addition to SMSC usual care or SMSC usual care alone (group B).

Patients allocated to the new treatment strategy have 6-monthly sNfL monitoring including 6-monthly blood draws and communication of sNfL values to treating physicians in addition to their usual care within the SMSC.

The monitoring of sNfL requires a 6-monthly blood draw. In practice, for the duration of the trial, patients allocated to group A who came to the SMSC every 12 months are asked to now come in-between their yearly visits (i.e., at 6-month) for an additional blood draw to monitor sNfL. For patients allocated to group A who came to the SMSC every 6 months anyway, the blood draw is always performed as part of the SMSC usual care, and no additional blood for the purpose of the trial will be taken. One serum aliquot per blood draw is used to measure sNfL centrally at the Clinical Neuroimmunology Laboratory, Department of Biomedicine in Basel, Switzerland. Two sNfL assays are used: single molecule array (Simoa, Quanterix, USA) and Cobas (Roche, CH) technologies. Percentiles and z scores normalized sNfL values (14) are reported to the clinicians within 14 days from blood sampling.

The SMSC treating physicians receive 6-monthly sNfL values for all patients allocated to the intervention arm and at a maximum 10 days after the patient’s visit. If and how the physician and patient acts upon the sNfL value is beyond the scope of this trial. To facilitate the implementation of sNfL in treatment decision-making, we have established treatment decision algorithms that integrated sNfL in addition to usual care assessments for the most common clinical scenarios (Appendix 2). Those algorithms were decided upon by consensus among experts in the field and patient consultant using a modified Delphi approach (31,32).

### Usual care comparator (group B)

Patients allocated to the usual care comparator (group B) continue with their usual care in the SMSC, including their usual 6 to 12 monthly visits.

Patients who do not consent to be randomly allocated and participate in the MultiSCRIPT-Cycle 1 trial will continue their SMSC usual care and will not be exposed to any influence of the study.

This group of patients will allow to explore the external validity of the randomized trial results using only the SMSC routinely collected data (for example by exploring differences of characteristics of patients in MultiSCRIPT-Cycle 1 and the other cohort patients), but will not be used to determine intervention effects (33).

### Participant timeline

The recruitment period will last 2 years and the trial conduct until the primary analysis will last 3 years, but participants will be followed until the end of the close-out phase for an additional 6 months (Figure 1). The expected duration of participants’ follow-up will range from 1.5 year to 3.5 years depending on their timepoint of recruitment. Table 1 illustrated study assessment timeline for individual participants.

**Figure 1:**
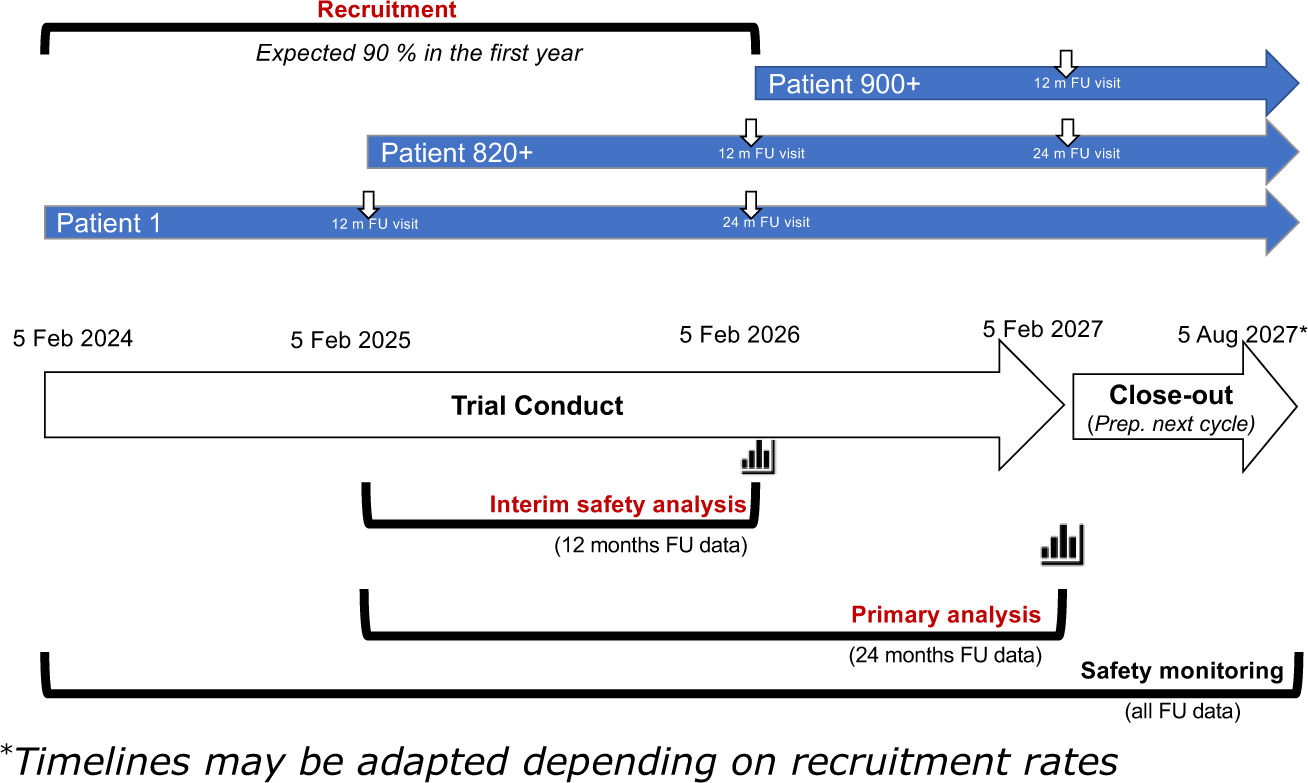
MultiSCRIPT Cycle 1 timeline.

**Table 1:** Study assessment timeline.

|  | Pre-allocation | Allocation | Post-allocation follow-up visits |  |  |  | Post-allocation Close-Out |  |  |
| --- | --- | --- | --- | --- | --- | --- | --- | --- | --- |
| Visit | - | V1 | V2 | V3 | V6 | V7 | V8 | V9 | V10 |
| Time | M0 (-14d) | M0 | M6 (±45d) | M12 (±45d) | M18 (±45d) | M 24 (±45d) | M30 (±45d) | M36 (±45d) | M42 (±45d) |
| **Recruitment** |  |  |  |  |  |  |  |  |  |
| Eligibility screening | X |  |  |  |  |  |  |  |  |
| Informed consent |  | X |  |  |  |  |  |  |  |
| **Randomization** |  | **X** |  |  |  |  |  |  |  |
| Baseline characteristics* | X | X |  |  |  |  |  |  |  |
| **Intervention** |  | **X** | **X** | **X** | **X** | **X** | **X** | **X** | **X** |
| **Primary outcomes** |  |  |  |  |  |  |  |  |  |
| MSQoL-54* |  | X |  |  |  | X |  |  |  |
| EDA3* |  | X |  |  |  | X |  |  |  |
| **Secondary outcomes** |  |  |  |  |  |  |  |  |  |
| MSQoL-54* |  | X |  | X |  | X |  |  |  |
| EDA3* |  | X |  | X |  | X |  |  |  |
| Relapses* |  | X |  | X |  | X |  |  |  |
| EDSS* |  | X |  | X |  | X |  |  |  |
| Lesions* |  | X |  | X |  | X |  |  |  |
| Amount of DMT*† |  |  |  | X |  | X |  |  |  |
| SAE |  | X | X | X | X | X | X | X | X |
| Indicators of harm* |  | X | (X) | X | (X) | X | (X) | X | (X) |
| - Relapses |  | x | (x) | x | (x) | x | (x) | x | (x) |
| - EDSS |  | x | (x) | x | (x) | x | (x) | x | (x) |
| - Opportunistic infections |  | x | (x) | x | (x) | x | (x) | x | (x) |
| **Other outcomes** |  |  |  |  |  |  |  |  |  |
| EQ-5D-5L* |  | X |  | X |  | X |  |  |  |
| Other PROs: |  |  |  |  |  |  |  |  |  |
| - Professional activity* |  | X |  | X |  | X |  |  |  |
| - Healthcare utilization - Hospitalization* |  | X |  | X |  | X |  |  |  |

\* Routinely collected data within the SMSC; (X) only when it coincides with a SMSC visit
† Defined as the daily dose following WHO recommendations

### Sample size

We plan to recruit all eligible patients from the SMSC and aim to recruit 915 patients. This estimation was based on number of eligible persons with RRMS in the SMSC across all 8 centres and assuming an 80% acceptance rate. We further estimate that the primary analysis will include 824 patients with 24-month follow-up data available. This was based on the assumption that we will recruit 90% of our target sample size the first year of recruitment.

Based on the data of the SMSC (date of last analysis: 15 October 2021), we assume 52% patients under usual care without sNfL biomarker monitoring will have an EDA3 during 24-month follow-up. We assume a relative risk reduction of 25% of EDA3 with sNfL biomarker monitoring compared to usual care to be a minimal important difference (MID). A sample size of 824 patients would have 93% power to detect the MID.

We are not aware of an established MID for MSQoL-54. We assume a difference of 0.2 (Hedges g) as a MID for the MSQoL-54, considering current guidelines for health-technology assessment and reimbursement decisions on quality of life assessments (34). Using a systematic search for trials using quality of life outcomes, we identified two recent trials (SUNBEAM (35) and RADIANCE (36)) using the MSQoL-54 instrument in a population that had a higher disease activity compared to the SMSC population. We used a conservative approach assuming a SD=20 based on the pooled standard deviation at baseline in the SUNBEAM (35) and RADIANCE (36) trials, an intra-patient correlation (baseline and 24m follow-up) of 0.8 and a correlation of the composite scores of 0.5 (r=0.66 reported by Vickrey et al (28)), which shows that a sample size of 824 patients in MultiSCRIPT-Cycle 1 would have > 95% power to detect the MID of 0.2 or a difference in 4 points on the MSQoL-54 total score. The power for 3 points would be 91% in the primary analysis. Given the pragmatic approach of MultiSCRIPT-Cycle 1 with a probably more heterogeneous study population than the more explanatory SUNBEAM and RADIANCE patient populations, a larger variability of quality of life may be plausible. A recently published survey in MS patients from the Netherlands, France, the United Kingdom, Spain, Germany and Italy, however, reported SD for the composites of 17.2 (physical) and 21.5 (mental) (29). However, even in a scenario with a 25% larger variability (SD of 25), the statistical power for detecting a difference of 4 points would be large with a sample size of 824 (94%).

The power calculations have been adjusted for multiple testing of the two primary outcomes.

### Recruitment and informed consent

We aim to recruit 915 participants which as of January 2024 represent 61% of the eligible SMSC participants with RRMS (n=1503). Participants are recruited by their SMSC physician during their usual care visit to the SMSC.

Making use of the existing SMSC database, eligible patients are identified ahead of their SMSC visit. Depending on the study site, the information sheet and consent form for the trial may be sent ahead of the patient’s visit. On the day of their SMSC visit, patients are invited to take part in MultiSCRIPT Cycle 1 by their SMSC treating physician. If they accept to sign the inform consent, patients are then randomized and their allocated group is directly communicated to them because patients allocated to the new treatment strategy need to know if they have to come back in 6 months.

We adopted proactive mitigation measures to address recruitment risks (37), including: (a) a dedicated ‘diagnosis’ of factors supporting and hindering recruitment informed by QuinteT Recruitment Intervention (QRI) methods, with a view to sharing good practice and training materials on how to recruit patients informed by QRI (38–40); (b) a recruitment log to record reasons for declining participation, informed by the SEAR process (41); (c) a quarterly newsletter will be submitted to all recruiting centers, reporting the progress and expected recruitment in all centers; and (d) online conferences (one per quarter in the first year of recruitment) and annual investigator meetings will be used to evaluate processes and share experiences.

## Assignment of treatment strategies

### Allocation

Participants are randomized in a 1:1 ratio to the trial groups stratified by study site. The randomization will be implemented in a centralized manner using the SMSC web-based electronic data capture implemented by RodanoTech, an electronic data capture (EDC) and data management services provider for the SMSC. Allocation concealment is ensured by the centralized and instant web-based randomization.

### Blinding procedures

MultiSCRIPT-Cycle 1 is a pragmatic trial that aims to provide evidence closely reflecting what happens in usual care. Therefore, being aware of the treatment and having a real-world assessment of treatment outcomes are part of the evaluated intervention within this pragmatic trial framework.

## Data collection and management

### Assessment and collection of outcomes

Most data related to outcome measures are routine collected within the SMSC and at a minimum on a yearly basis. For MultiSCRIPT-Cycle 1, the primary outcomes will be assessed at 24-months follow-up and secondary outcomes at 12- and 24-months follow-up. The additional data collected for the purpose of MultiSCRIPT are the sNfL measurements, and MRI related data based on radiological reports from the local centers.

### Retention and adherence

MultiSCRIPT-Cycle 1 uses a pragmatic approach, and therefore no specific steps are taken to maximize adherence for the purpose of the trial as this would create an artificial setting and deviate from usual care. Patients are followed within the SMSC and the SMSC usual care mimics routine care for patients with MS.

Treatment changes will be recorded but no mitigation action will be taken to enforce implementation of the treatment decision algorithms. It remains the treating physician and the patient’s choice to implement a treatment change based on sNfL as it would occur in usual care.

## Statistical methods

### Statistical analysis for primary and secondary outcomes

Superiority of a treatment strategy will be assumed when there is an improvement in at least one of the two primary outcomes, i.e., the strategy either reduces the risk of EDA3 *<u>or</u>* increases quality of life. Since there are two primary outcomes the significance level will be 2.5%.

### Primary Analyses

The primary analyses for the two primary outcomes will be conducted when 90% of the total number of participants (i.e. n= 824) have been recruited, randomized, and have at least 24 months follow-up data (expected to occur after 3 years of trial conduct). The primary analysis will follow the intention to treat (ITT).

To test the between-group difference 24 months after randomization regarding the proportion of patients experiencing at least one EDA3 event, the Pearson’s χ^2^-test will be applied. Odds ratio and 95% confidence intervals will be reported. To test the between-group difference in the MSQoL-54 scores 24 months after randomization, a linear regression model will be used. The model will be adjusted on the following covariate: baseline MSQoL-54 score. The score differences and 95% confidence intervals will be reported. To adjust for multiple testing of the two primary outcomes, the Bonferroni correction will be applied and the threshold for rejecting null hypotheses will be set at α = 2.5%.

### Secondary Analyses

The analysis of the primary outcomes will be repeated with 12-month follow-up data using all patients who were randomized and thus have a minimum of 12-month follow-up. In addition, each component of the EDA3 primary outcome (i.e., proportions of patients with at least one relapse, EDSS worsening, new/enlarging T2w lesion or T1w contrast enhancing lesion, and proportions of patients with at least two criteria of EDA3) and of the MSQoL-54 score (i.e., physical health composite summary and the mental health composite summary) will be assessed using Pearson’s χ^2^-test and linear regression models, respectively. Secondary outcomes will also be assessed at 24-month follow-up.

The amount of immunosuppressive drug treatment will be summarized using the defined daily dose as defined by the World Health Organization (WHO) (42) i.e., the assumed average maintenance dose per day for a drug used for its main indication in adults.

### Interim Harm analyses

An interim analysis will be conducted when 90% of the total number of participants (i.e. n= 824) have been recruited, randomized, and have at least 12 months follow-up data (expected to occur after 2 years of trial conduct). The rationale being that indicators of harms are collected at a minimum on a yearly basis (as some participants only come once a year to the SMSC) we therefore need a minimum of one year follow-up data. Using 12-month follow-up data, the interim analyses will focus of reviewing any SAEs related to blood draw and indicator of harms including mortality, harms related to immunosuppression (e.g., relevant infections) and any event related to occurrence of relapses and/or disability worsening in patients previously stable. The analysis will be conducted and reviewed in a blinded fashion by the Data Safety Monitoring Board (DSMB) made of study-independent experts, including a statistician and experts in trial methodology, ethics and/or care management in MS.

The DSMB will request unblinding of the data if any of the aforementioned outcomes exhibit statistically significant differences when comparing the two trial groups. We will consider that the causal pathway between the key intervention (i.e., monitoring and providing the sNfL information) and indicators of harms is complex and difficult to assess because sNfL is only one parameter among many that is considered in shared decision-making about treatment adaptation in usual care.

Interim analyses will be considered indicative and will not lead to an automatic early stopping of the trial. In case of major safety concerns, based on their expertise and review of the literature, the DSMB may recommend an early termination of the trial. Of note, “efficacy” will not be assessed in the interim analysis.

### Harm analysis

Incidence rates of SAEs and multiple occurrences of SAEs within individual patients will be reported separately and summarized and compared between treatment arms using the Pearson’s χ^2^-tests. Harm outcomes will be assessed until the end of the assessment cycle (i.e., up to 42-month follow-up).

### Additional analyses

Subgroup effects on the primary outcomes will be analyzed by interaction tests. To this end, EDA3 will be assessed in a logistic regression model and MSQoL-54 in a linear regression model. All subgroup analyses will be pre-specified in the final statistical analyses plan (SAP) which will be published before unblinding the data.

The key subgroups of interest are patients with low or high baseline risk of further EDA (secondary objective; section 5). In addition, we plan to analyze the following subgroups: age (<40 years vs. >40 years); Sex (male or female); Treatment-naive or previously treated for multiple sclerosis (with any DMT at any time before study enrolment); Number of relapses in the year before the study (≤1 relapse or >1 relapse); Number of relapses in the 2 years before the study (≤1, 2, or >2 relapses); Baseline disability (EDSS score 0–3.5 or >3.5); Number of contrast enhancing lesions at baseline (0 or ≥1); Number of T2w lesions (<3; 3 to 9; >9); and T2w lesion volume (≤3300 μL or >3300 μL).

We will consider the criteria as determined by the ICEMAN tool when interpreting subgroup effects and reporting the results (43).

In addition, to better understand the causal pathway that leads to the outcomes, we will also explore the outcomes in patients who have been treated as expected according to the treatment guidelines (e.g., patients with a relatively clear indication to escalation and subsequently escalated). In the same direction, we will explore the treatment decision (categorical: no (major) change, escalation or de-escalation) as a mediator in the causal pathway from the intervention to the outcomes, as we do not expect the intervention to have a direct causal effect on the outcomes, but an indirect effect through this mediator (complete mediation). For this, we will record each treatment changes. We will also explore the impact of lack of blinding using the centralized reading of MRI compared with local MRI reports.

Since such analyses rely on complex assumptions, are based on non-randomized comparisons, and are prone to time-dependent confounding bias, we will carefully prespecify such additional analyses consider them entirely as exploratory (44).

Finally, health economic analyses will be detailed in a specific health economic analysis plan that will be finalized before the primary analysis starts.

### Handling of missing data and drop-outs

Patients are already participating in the SMSC which include at a minimum yearly visit with an assessment of their EDA3 status (i.e., EDSS, relapses and MRI data are routinely collected), thus the occurrence of missing data should be minimal for the EDA3 primary outcome. MSQoL-54 is also part of the SMSC usual care and in addition to careful planning and conduct of the trial should also minimize the occurrence of missing data on the MSQoL-54 primary outcome. To test the robustness of our results to missing data, various imputation methods will be implemented, and a detailed description of imputation methods will be provided in the SAP. In the unlikely event that missing data exceeds 40%, we will only report the complete cases analysis (45) and we will interpret it only exploratory.

### Dissemination plans

Statistical code will be made available on GitHub while the patient-level data set will be made available upon request to the primary investigator. Results will be published in an open-access peer-reviewed medical journal.

## Discussion

MS is the most frequent non-traumatic cause of disability in young adults and MS patients have a lower health related quality of life compared with the general population but also compared with other chronic diseases (46,47). An estimated 15,000 persons are living with MS in Switzerland (48) of whom >1800 are included in the SMSC with a median follow-up of 6.3 years, as of January 2024. Patient care in the SMSC reflects the usual care of MS patients nationwide.

Within the SMSC, almost half of the patients with RRMS receive DMTs known for their high efficacy but with uncertainties on the long-term exposure to these treatments. A significant number (over 200) of patients in the SMSC also remain untreated and potentially at risk of disease activity. Improvement of patients’ quality of life by providing patients and physicians with information that may reduce the uncertainty and enable a biomarker guided de-escalation when applicable would be highly relevant and a significant step towards a more personalized approach to MS treatment and care management. This may reduce the treatment burden and the risk for potential severe side effects associated with highly effective DMT. On the other hand, there may be a better basis for informed decision-making and better informed, earlier and/or more sensitive biomarker-guided escalation that may lead to fewer patients with evidence of disease activity. Better and adequate monitoring of treatment response would also reduce the costs generated by over- or under treating patients. Moreover, a continuous sNfL monitoring may improve patients’ quality of life by providing an additional sense of security in a disease which is largely unpredictable.

The pragmatic attitude of MultiSCRIPT being fully embedded in the SMSC usual care with minimal disruption to patient care, leveraging routinely collected data and aiming to generate evidence to inform decisions lays the foundation for creating a learning healthcare system. A learning system whereby accumulating data will enable the continuous generation of new hypotheses on how treatment and care strategies can be further personalized to treat patients as little as possible but as much as necessary at the right time, i.e., ensuring no evidence of disease activity (with more sensitive biomarkers such as for example sNfL), while achieving better patient-relevant outcomes, and improved care by better informing shared decision-making.

## Trial status

The first patient has been included on 5 February 2024. At the submission of this manuscript on 22 March 2024, 105 patients have been included and randomized and recruitment is ongoing.

## Data Availability

All data produced in the present studywill be available upon reasonable request to the authors

## Declarations

## Acknowledgments

We would like to thank all study coordinators and nurses involved in the SMSC as well as persons with MS participating in the SMSC. We would also like to thank the patient consultants who provided feedback on the protocol proposal.

## Authors contributions

Authors contributions according to the CRedIT author statement:

1. Conceptualization: PJ, LK, JK, LGH, ÖY
2. Data Curation: ./.
3. Formal Analysis: PJ, PB, SS, LGH
4. Funding Acquisition: CZ, AS, JK, LGH, ÖY
5. Investigation (trial-ongoing): CZ, AO, LD, AM, CG, JO, TD, LA, OF, PR, PL, SM, CP, RH, GD, CG, JK, ÖY
6. Methodology: PJ, SS, LR, MS, MJL, LGH
7. Project Administration: PJ
8. Resources: ./.
9. Software: ./.
10. Supervision: JK, LGH, ÖY
11. Validation: ./.
12. Visualization: ./.
13. Writing – Original Draft: PJ, LGH
14. Writing – Review & Editing: all authors

## Ethics approval and consent to participate

The Ethikkommission Nordwest- und Zentralschweiz (EKNZ) reviewed and approved the protocol (BASEC 2023-01578). Written, informed consent to participate will be obtained from all participants.

## Consent for Publication

Not applicable

## Appendix 1: PRECIS-2 assessment

PRECIS-2 scoring results are (pragmatic attitudes are reflected by a maximum of 5 points, while explanatory characteristics by lower scores) - Eligibility 5 (broad eligibility criteria with only one exclusion criterion); Recruitment 5 (patients will be recruited during their usual care yearly visit); Setting 5 (embedded in the SMSC usual care and 7 centers from the cohort will be recruiting); Organization 5 (make use of the already existing SMSC organization in place); Flexibility – delivery 5 (treatment guidelines pre-specified but as any guidelines, it will be the physicians and patients’ decision to follow them); Flexibility – Adherence 5 (no mitigation measures will be taken to increase the use of diagnostic information); Follow-up 5 (number of visits and routine assessment increased in intervention group, but part of the biomarker monitoring intervention; no change in control group); Primary Outcome 5 (decision-relevant and patient reported outcome); Primary analysis 5 (intention to treat).

## Appendix 2: Treatment decision algorithms

The following treatment decision algorithms are intended to serve as treatment decision aids for the clinical application of serum neurofilament light chain values within MultiSCRIPT-Cycle 1 to guide escalation and de-escalation of disease modifying therapies (DMTs) in patient with relapse-remitting multiple sclerosis (RRMS). They represent a minimal set of treatment decision algorithms that experts have reach a certain level of agreement on.

The proposed treatment decision algorithms are not meant to cover all unique cases that physicians might encounter in the clinics. We acknowledge that treatment modifications related to pregnancies, treatment intolerances, comorbidities and other causes may not be captured here. The proposed algorithms are not binding and do not intend to supplant patients’ and physicians’ preferences.

Physician at their discretion may pursue any *additional clinical and/or clinical assessment* as per usual care to inform the decision.

The treatment decision algorithms rely on the following definitions:

**Usual care arm:** Consider escalation, if there is evidence of disease activity (clinically or MRI-activity)

**NEDA:** no evidence of disease activity

**NEDA2:** no relapse, no EDSS worsening

**NEDA3:** no relapse, no EDSS worsening, no MRI activity

**EDSS worsening:** defined as an increase of ≥1.5 points from an EDSS of 0, ≥1.0 point from an EDSS of 1.0-5.0 or ≥0.5 point from an EDSS ≥5.5

**MRI activity:** Any unequivocal new or enlarging T2w lesion (***after adequate re-baselining***) or contrast enhancement on T1w images on brain or spinal cord MRI according to the consensus between local neuroradiologist and treating neurologist.

**Evidence of disease activity based on sNfL** is defined using sNfL>90th percentile (<u>Benkert et al. Lancet</u> <u>Neurology 2022</u>) and ***once other potential causes of high sNfL have been excluded*** (e.g. trauma, stroke, relevant sports-related head injury, at least medium severe renal failure (GFR < 60 mL/min/1.73 m2), suboptimally treated diabetes mellitus or any other concomitant disease that may lead to relevant neuroaxonal damage)

| DMT CLASSIFICATION | Delphi results |
| --- | --- |
| **Low efficacy:** glatiramer acetate – interferon beta – teriflunomide | 97% Strong consensus |
| **Medium efficacy:** fumarates – S1P modulators – cladribine | 83% Broad consensus |
| **High efficacy:** alemtuzumab – anti B cell therapy – natalizumab | 100% Full agreement |

| ESCALATION TREATMENT DECISION ALGORITHMS |  | Delphi results |
| --- | --- | --- |
| **TREATMENT INITIATION FROM UNTREATED** |  |  |
| If your patient is currently **untreated** and has high sNfL (>90th percentile) jointly consider with your patient to <u>initiate DMT</u> if your patient has.... |  |  |
| ▪ **NEDA2 plus MRI activity with at least 2** new/enlarging T2w lesions or contrast enhancing T1w lesions |  | 97% Strong consensus |
| ▪ **NEDA2 plus MRI activity with at least 1** new/enlarging T2w lesion or contrast enhancing T1w lesion |  | 93% Strong consensus |
| ▪ **NEDA 3** |  | 80% Broad consensus |
| **ESCALATION FROM LOW DMT** |  |  |
| If your patient is currently receiving a **low efficacy DMT** for at least 9 months and has high sNfL (>90th percentile) jointly consider with your patient to escalate to <u>medium or high efficacy DMT</u> if your patient has..... |  |  |
| ▪ **NEDA2 plus MRI activity with at least 2** unequivocal new/enlarging T2w lesions or contrast enhancing T1w lesions |  | 97% Strong consensus |
| ▪ **NEDA2 plus MRI activity with at least 1** unequivocal new/enlarging T2w lesion or contrast enhancing T1w lesion |  | 97% Strong consensus |
| ▪ **NEDA3** |  | 67% Moderate consensus |
| **ESCALATION FROM MEDIUM DMT** |  |  |
| If your patient is currently receiving a **medium efficacy DMT** for at least 9 months and has high sNfL (>90th percentile) jointly consider with your patient to escalate to <u>high efficacy DMT</u> if your patient has... |  |  |
| ▪ **NEDA2 plus MRI activity with at least 2** unequivocal new/enlarging T2w lesions or contrast enhancing T1w lesions |  | 100% Full agreement |
| ▪ **NEDA2 plus MRI activity with at least 1** unequivocal new/enlarging T2w lesion or contrast enhancing T1w lesion |  | 93% Broad consensus |
| ▪ **NEDA3** |  | 67% Moderate consensus |

| HORIZONTAL SWITCH TREATMENT DECISION ALGORITHMS | Delphi results |
| --- | --- |
| **HORIZONTAL SWITCH FROM NATALIZUMAB** |  |
| If your patient is currently receiving **natalizumab** and has high sNfL (>90thpercentile) jointly consider with your patient to <u>switch to a different mode of action high efficacy DMT</u> if your patient has... |  |
| ▪ **at least 9 months** natalizumab plus **NEDA2** plus **MRI activity with at least 2** unequivocal new/enlarging T2w lesions or contrast enhancing T1w lesions | 97% Strong consensus |
| ▪ **at least 9 months** natalizumab plus **NEDA2** plus **MRI activity with at least 1** unequivocal new/enlarging T2w lesion or contrast enhancing T1w lesion | 84% Broad consensus |
| ▪ **at least 12 months** natalizumab plus **NEDA3** | 54% Moderate consensus |
| **HORIZONTAL SWITCH FROM ANTI-B CELL THERAPY** |  |
| If your patient is currently receiving **anti-B cell therapy** and has high sNfL (>90thpercentile) jointly consider with your patient to <u>switch to a different mode of action high efficacy DMT</u> if your patient.... |  |
| ▪ **at least 9 months** anti-B cell therapy plus **NEDA2** plus **MRI activity with at least 2** unequivocal new/enlarging T2w lesions or contrast enhancing T1w lesions | 93% Broad consensus |
| ▪ **at least 9 months** anti-B cell therapy plus **MRI activity** plus **confirmed EDSS worsening or relapse** | 93% Broad consensus |
| ▪ **at least 12 months** anti-B cell therapy plus **NEDA2** plus **MRI activity with at least 1** unequivocal new/enlarging T2w lesion or contrast enhancing T1w lesion | 92% Broad consensus |
| ▪ **at least 12 months** anti-B cell plus **no MRI activity but relapse** | 83% Broad consensus |
| ▪ **at least 12 months** anti-B cell plus **no MRI activity but confirmed EDSS worsening** | 71% Moderate consensus |
| **HORIZONTAL SWITCH FROM CLADRIBINE** |  |
| If your patient is received the second cycle **cladribine** and has high sNfL (>90thpercentile) jointly consider with your patient to switch to <u>a different mode of action or regimen strategy of DMT</u> if your patient has.... |  |
| ▪ **at least 6 months** after the second cycle plus **NEDA2** plus **MRI activity with at least 2** unequivocal new/enlarging T2w lesions or contrast enhancing T1w lesions | 97% Strong consensus |
| ▪ **at least 12 months** after the second cycle plus **NEDA3** | 83% Broad consensus |
| ▪ **at least 6 months** after the second cycle plus **NEDA2** plus **MRI activity with at least 1** unequivocal new/enlarging T2w lesion or contrast enhancing T1w lesion | 81% Broad consensus |

| DE-ESCALATION TREATMENT DECISION ALGORITHMS | Delphi results |
| --- | --- |
| **DE-ESCALATION FROM ANTI-B CELL THERAPY** |  |
| If your patient is currently receiving **anti-B cell therapy** for at **least 2 years**, has **NEDA3 for the past 2 years** and has normal sNfL (< 80th percentile) jointly consider with your patient to perform 6-monthly cMRI and 6-monthly sNfL measurement and de-escalate by **extending treatment interval up to 12 months (Ocrevus, Rituximab) or 8 weeks (Kesimpta)** | 74% Moderate consensus |
| **DE-ESCALATION FROM FUMARATES** |  |
| If your patient **is >60 years old**, currently receiving **fumarates** for at **least 5 years**, has **NEDA3 for the past 5 years, pre-treatment activity was low** (e.g., less than 1 relapse per year) and has normal sNfL (<80th percentile) jointly consider with your patient to perform 6-monthly cMRI and 6-monthly sNfL measurement and **stopping fumarates** | 74% Moderate consensus |
| **DE-ESCALATION FROM LOW EFFICACY DMT** |  |
| If the patient wishes to stop treatment and is **>60 years old**, currently receiving **low efficacy DMT** for at **least 5 years**, has **NEDA3 for the past 5 years** and has normal sNfL (< 80th percentile) jointly consider with your patient to perform 6-monthly cMRI and 6-monthly sNfL measurement and **stopping DMT treatment** | 87% Broad consensus |

